# Patient-level micro-simulation model for evaluating the future potential cost–effectiveness of pharmacy-based interventions in the control and management of diabetes-related complications in Canada

**DOI:** 10.1101/2020.03.10.20033597

**Authors:** Mohsen Yaghoubi, Kerry Mansell, Hassanali Vatanparast, Wu Zeng, Mehdi Javanbakht, Marwa Farag

## Abstract

**Background:** The increased risk of complications among diabetes patients poses a serious threat to population health. Pharmacy-based interventions can decrease the burden of diabetes and its related complications. This study evaluates the cost-effectiveness of pharmacy-based interventions and offers insights on the practicality of their adoption by health practitioners.

**Methods:** We developed population-based micro-simulation model using 2,931 patients with diabetes in Canada. We used the risk equations on the UK Prospective Diabetes Study (UKPDS) to estimate the incidence and mortality of four of the most common diabetes-related complications (heart failure, stroke, amputation, and blindness). We extrapolated the potential effects of pharmacy interventions on reducing time-varying risk factors for diabetes complications. Cost was quantified as the annual cost of complications; and, the cost associated with pharmacy-based interventions. The final outcomes were the incremental costs per quality-adjusted life years (QALY) gained. Both deterministic and probabilistic sensitivity analysis were conducted to examine the robustness of the ratio.

**Result:** Pharmacy-based interventions could prevent 155 preventable deaths, 159 strokes, 29 cases of blindness, 24 amputations, and 19 heart failures across the lifetime of 2,931 patients. In addition, an estimated 953 QALYs (0.32 per patient) would be gained among the intervention group. Per QALY, the incremental discounted cost is $3,928, suggesting that pharmacy-based interventions are likely cost-effective compared to usual care. At an ICER threshold of $50,000, over 92% of the simulation remains cost-effective.

**Conclusion:** Pharmacist-based interventions targeted at addressing the development of diabetes-related complications among Canadian patients have the potential to offer a cost-effective strategy.

## Introduction

Diabetes mellitus (DM) is one of the most prevalent non-communicable diseases worldwide, and poses a major problem to the 21^st^ century health system (1, 2). Perhaps most threatening about DM is its unprecedented growth: In less than three decades, the number of adults diagnosed with diabetes has doubled (2). This translates to over 400 million people living with diabetes today, and an estimated 642 million who will live with diabetes by 2040 (3). In many developed countries, diabetes is a leading cause of death, and was the culprit behind at least 1.3 million deaths in 2013 alone (4). Diabetes is also associated with a number of complications, including: cardiovascular disease, nephropathy, neuropathy, amputations, and blindness (5-8). These complications lead to premature death, reduce individuals’ quality of life, and place a heavy global economic burden on the whole of society to the tune of US$548 billion each year (3, 5-8). This cost is not limited to the health care system, but also includes indirect costs incurred by loss of productivity resulting from disability and/or premature death (9). Unfortunately, the prognosis in Canada is not much better. In comparison to the general population, diabetes patients are three times more likely to be hospitalized for cardiovascular disease, twelve times more likely to be hospitalized for end-stage renal disease, and twenty times more likely to be hospitalized for a non-traumatic lower limb amputation (11, 12). This means that, of the Canadian healthcare dollars being expended on diabetes patients, eighty percent are incurred as a result of diabetes-related complications (10). However, there is an opportunity to reduce this expenditure through the adoption of diabetes management interventions such as pharmacy-based ones. Pharmacy-based interventions include a wide range of services, all with the common aim of giving diabetes patients greater control and management over their disease. Common examples of pharmacy-based interventions include: consultations with pharmacists; patient education about self-monitoring and self-management techniques; preventive programming that emphasizes lifestyle modifications; reminders about annual physical examinations; assistance with adherence to medication; patient education about the correct use of insulin, anti-hyperglycemic medications and oral hypoglycemic agents; and, programming that increases patients’ awareness about effective diabetes management (13-17). Recent reviews of pharmacy-based interventions have demonstrated that they have a positive impact on clinical outcomes (13-17). These findings are promising, especially because they suggest that pharmacy-based interventions could reduce diabetes-related complications, morbidity and mortality (13-17). Rising health care costs, limitations on available health care resources, and debates over the comparative effectiveness of diabetes management strategies has led to an increased interest in developing analytic models that can evaluate the future potential cost-effectiveness of such intervention. These models compliment clinical trials, which typically provide data on intermediate outcomes like HbA1c, SBP, and LDL. Data from clinical trials can then be used to populate analytic models, and provides a basis for predicting long-term health outcomes, like life-years saved or QALYs gained.

This study aims to evaluate the future potential cost effectiveness of pharmacy-based interventions for diabetes management in Canada. In order to conduct this evaluation, we estimate life time outcome and quality-adjusted life expectancy among diabetes patients who experience diabetes-related complications (heart failure, stroke, amputation, and blindness) or diabetes-related death.

## Methods

### Model Overview

We evaluated the future potential cost-effectiveness of pharmacy-based interventions for patients with diabetes compared to usual care. Intermediate outcome of intervention were modeled as reduction of hemoglobin A1C (HbA1c) level, body mass Index (BMI), systolic blood pressure (SBP), and low-density lipoproteins (LDL) as most important risk factors of four common diabetes-related complications including heart failure, stroke, amputation and blindness. Cost was quantified as the annual cost of heart failure, stroke, amputation and/or blindness among diabetes patients; and, the cost associated with pharmacy-based interventions borne by society. To fully capture the effect of the intervention, we extrapolated the potential effects of intervention in relation to cost, health outcomes, and health-related quality of life (HRQoL) over the next 50 years by calculating the incremental cost per QALY gained of pharmacy-based intervention versus usual care in base case scenario. This model considered both costs and health effects, which were adjusted by discount rate of 3% according to the Canadian Agency for Drugs and Technologies in Health (CADTH) guideline (18). Both deterministic and Monte Carlo probabilistic sensitivity analysis were used to estimate uncertainty around results. Detail of model overview is shown in ***Figure (1)***.

**Figure 1).**
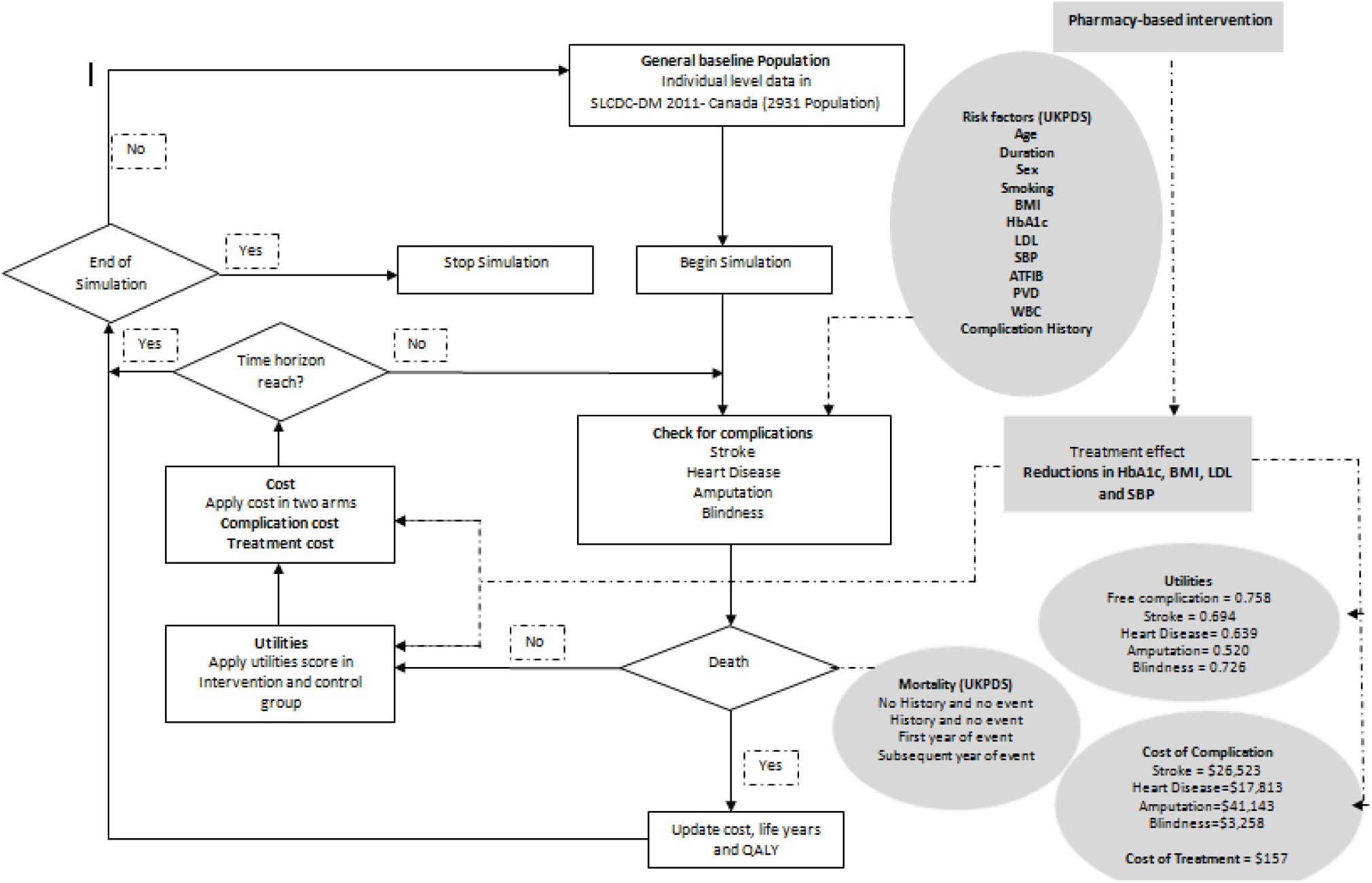
Flow chart of simulation.

### Model Structure

Using Anylogic software package 8.2.3, a hybrid simulation model was developed, which included agent-based and system dynamics. Agent-based modeling (ABM) enables to simulate more complex interactions and processes associated with chronic disease. Thus, this technique is very suitable for incorporating individuals with different risk factors and health behavior characteristic and evaluating the impact of adjusting risk factors on the better control and management of diabetes (19).We captured four major diabetes-related complications progresses among patients with heterogeneous characteristics through state-transition formalism as state charts in ABM model. Also, we used a system-dynamic approach to estimate accumulated costs and QALYs over time ***(Figure A1-A2 in appendix)***.

The simulation model mimicked a multistage study and was populated by data from the Survey on Living with Chronic Diseases in Canada (SLCDC). This data was used to build an individual-level micro-simulation model predictive of diabetes-related complications (heart failure, stroke, blindness, and amputation) and death, and the associated health care cost and QALYs in the presence and absence of pharmacist-based intervention. Within the model, a set of attributes known to be associated with diabetes-related complications was assigned to each person. The attributes were also subject to a set of rules (i.e. transition probabilities) and states reward (i.e. cost and utility). All parameters are shown in ***Table (1)***.

### Data source

#### Diabetes Risk Factors

To estimate a diabetes patient’s progression from complication free to a complication ridden state, we used the UK Prospective Diabetes Study (UKPDS) outcome model (20). This model was developed using data from 5,102 patients followed over a 20-year trial period, and 4,031 survivors followed over a 10-year post-trial monitoring period. The UKPDS model allowed us to estimate the main risk equations for developing the four diabetes–related complications of interest in our study: heart failure, stroke, amputation and blindness. It also allowed us to derive parametric proportional hazard models predictive of absolute risk factors of diabetes-related complications, including: age, sex, duration of diabetes, smoking status, body mass index (BMI), HbA1c, SBP, LDL, high density lipoprotein cholesterol (HDL), heart rate, presences of micro- or macro-albuminuria (MICLAB), atrial fibrillation (ATFIB), peripheral vascular disease (PVD), white blood cell count (WBC), amputation history, heart failure history, stroke history, blindness history, renal disease history, and ulcer foot history. UKPDS model used a Weibull proportional hazards regression to calculate the occurrence of the composite outcome, which combined both fatal and non-fatal events (20). We extracted baseline characteristics from the 2,931 diabetes patients included in the SLCDC survey 2011 ***(Table 1)*** and risk equations for each of the baseline variables were taken from the UKPDS. This allowed us to estimate transition probabilities of developing diabetes-related complications ***(See Appendix I and Table A1)***. For some of the risk questions which the corresponding baseline variables were not available in the SLCDC survey, we estimated baseline variables based on age and sex specific.

**Table (1).**
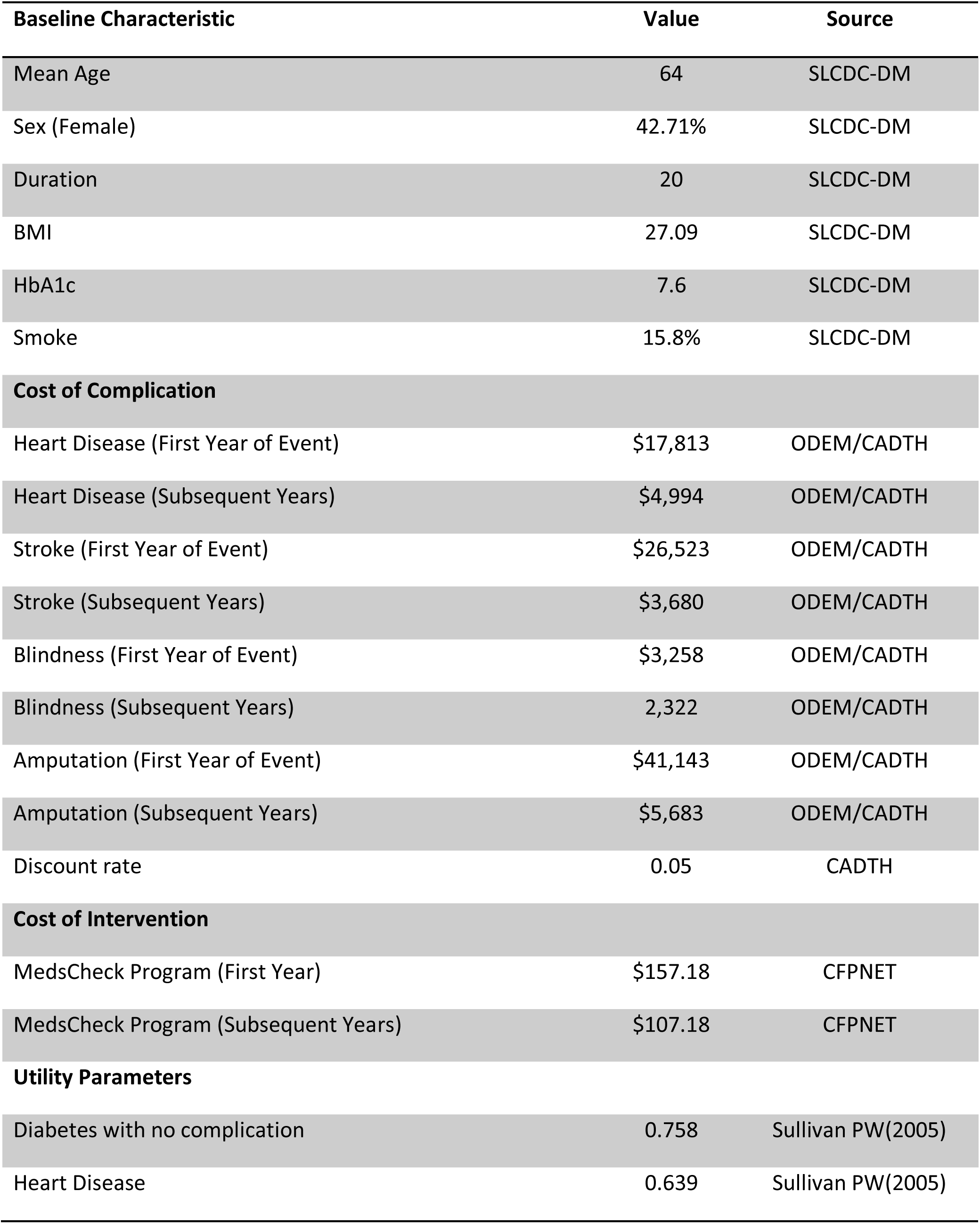
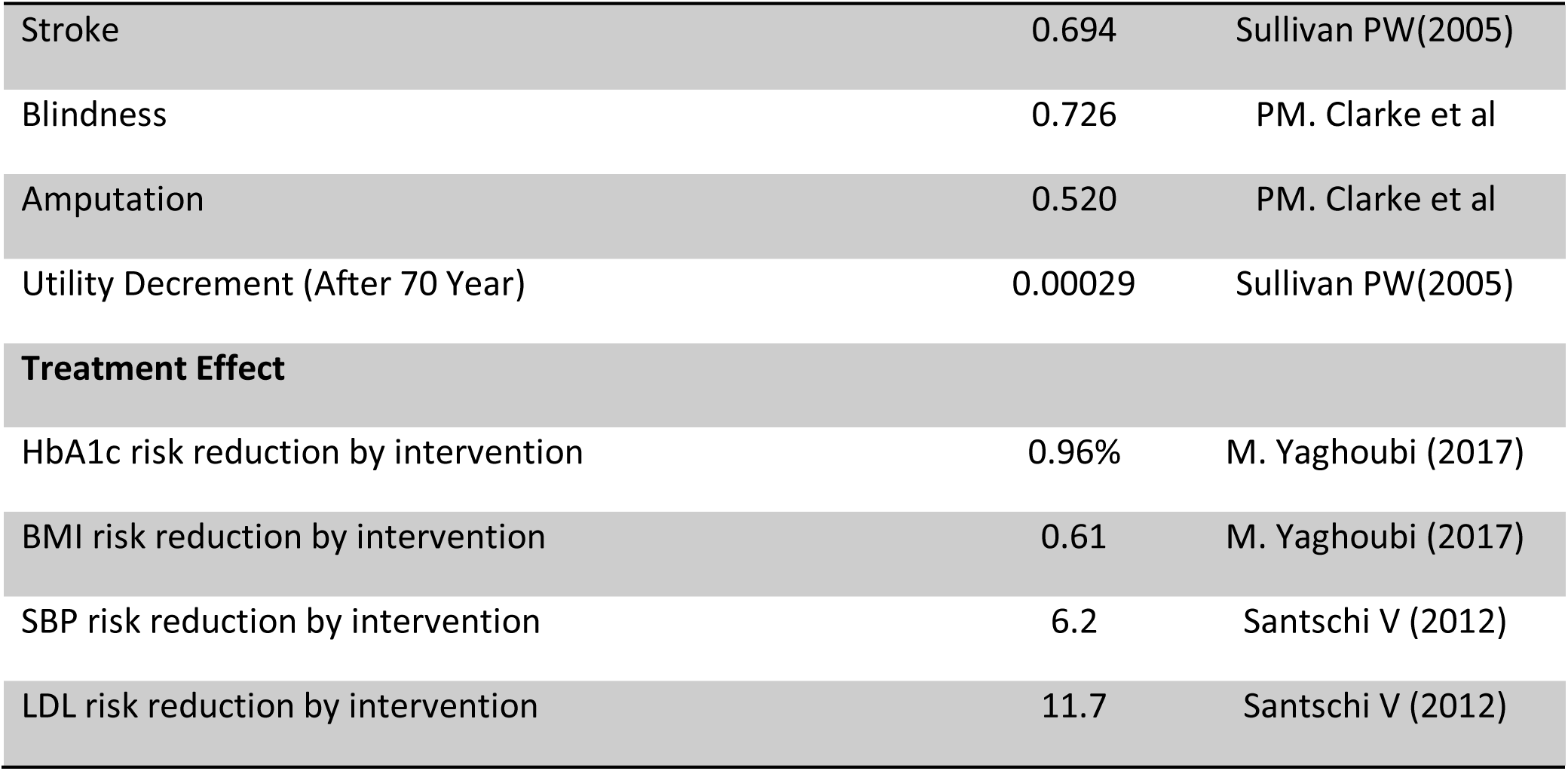
Microsimulation Model Parameters.

### Mortality

To estimate progression from a complication-free state to a state where at least one of the four major complications or death have occurred, our analysis considered four equations for calculating risk of mortality based on the UKPDS model. The first equation estimates the probability of death in the first year following an occurrence of heart failure, stroke, amputation or blindness, based on a logistic regression. Similarly, the second equation is based on logistic regression. It estimates the risk of diabetes-related mortality among patients with a history of any one of these complications in all subsequent years. The third equation is based on multivariate Gompertz proportional hazards survival models, and estimates death among diabetes patients without any history of diabetes-related complications. In this equation, death is the result of a cause unrelated to diabetes. The fourth equation is slightly more nuanced, and estimates death among diabetes patients without complication who had a history of co-morbidities (20). Transition probabilities of death were calculated from these equations based on logistic regression and Gompertz proportional hazards model ***(See Appendix II and Table A2)***.

### Treatment Effect

The impact that pharmacy-based interventions have on four major risk factors associated with diabetes-related complications was extracted from recent systematic reviews and meta-analyses. The four major risk factors include: HbA1c, BMI, SBP, and LDL.

#### Hemoglobin (HbA1c)

From baseline to the last follow-up 12 months later, the standardized absolute mean difference in the reduction of HbA1c was significantly more favorable in the pharmacy-based intervention group, then in the control group (0.96%; 95% CI 0.71:1.22, P<0.001) (13).

#### Body Mass Index (BMI)

From baseline to the last follow-up 12 months later, the standardized absolute mean difference in the reduction of BMI units was 0.61 (95% CI 0.20: 1.03, P=0.000) in favor of the pharmacy-based intervention group (13).

#### Systolic Blood Pressure (SBP)

In comparison to the control group, there were significant reductions in SBP among diabetes patients in the pharmacy-based intervention group after a 12-month period (−6.2 mmHg [95% CI −7.8 to −4.6]) (17).

#### Low Density Lipoprotein (LDL) Cholesterol

In comparison to the control group, there were significant reductions in LDL cholesterol among diabetes patients in the pharmacy-based intervention group after a 12-month period (−11.7 mg/dL [−15.8 to −7.6]) (17).

### Health Utilities

We assumed that utility values derived from the American population are relevant to the Canadian population. We quantified HRQoL for a set of health states of interest based on American catalogue of EQ-5D utility values (21). Health states of interest include: following a stroke, following heart failure, after the age of 70 years. Following a stroke, the resulting utility was 0.694. Following heart failure, the resulting utility was 0.636. After the age of 70, a utility decrement of 0.00029 per year was applied to all years.

Under the assumption that utility values derived from the United Kingdom population are relevant to Canada, the UKPDS outcome model was used to estimate HRQoL for a set of health states of interest. Health states of interest include following an amputation, and following blindness. Following amputation, a utility decrement of 0.520 was applied. Following blindness, a utility decrement of 0.726 was applied. A weight of zero was assigned to death. If applicable to incorporate the effect of concurrent complications, a multiplicative method approach was applied (21).

### Cost of treating diabetes-related complications

Health care resource utilization and their costs associated with the management of diabetes-related complications were extracted from the Ontario Ministry of Health and Long-Term Care (22). Total costs of patients with diabetes and patients with complications were quantified in terms of hospitalizations, outpatient visits, emergency visits, home care and long-term management costs. Costs were inflated to the value of the 2016 Canadian dollar using the health component of the Canadian Consumer Price Index. On average, the annual cost for patients without diabetes-related complications was $2,075 (22). The estimated costs for each of the four diabetes-related complications in the first year they occur, and for all subsequent years are shown in ***Table (1)***.

### Intervention Costs

To estimate the cost of pharmacy-based interventions, costs incurred by the implementation of the MedsCheck program served as a benchmark. This program is a pharmacy-based intervention targeted at diabetes patients in Canada (23). Based on the current fee schedule of the MedsCheck program in Alberta, the unit cost of the first annual consultation was determined to be $75 CAD, the unit cost of each subsequent consultation within that year was $25 CAD, and the unit cost of long-term follow-up was $75 CAD (23).

The indirect costs associated with wait-times and travelling related to pharmacy-based interventions were added to the model. Based on the MedsCheck program, the total time lost for pharmacy consultation was 2 minutes for waiting and 20 minutes for the duration of the consultation (24). Based on these estimates, we calculated an opportunity cost. The total time lost was multiplied by the number of pharmacy consultations in one year assuming that the entire study population would be the recipient of pharmacy-based interventions. This number was then multiplied by $24.96 CAD or the average wage per hour in Canada (25).

Based on data from Geographic Accessibility of Community Pharmacies, the cost of travelling to a pharmacy was estimated using the average travel time among patients who visited a pharmacist (26), and the mean fuel cost per kilometer (km) of $0.12/km CAD (27).

### Cost-effectiveness analysis

An incremental analysis combined the joint estimates of costs and effects across the baseline scenario and the intervention scenario. The result of this analysis yielded the point estimate of the mean ICER. The ICER was calculated as the difference in costs between baseline and intervention divided by the difference in effects (i.e. QALYs) between baseline and intervention.

Measures of variance for the joint incremental costs and effects were obtained using Monte Carlo simulation and presented graphically using the cost-effectiveness plane. In order to convert health outcome (QALYs) to common metric as dollar, the net monetary benefit (NMB) was calculated. The NMB is equal to the incremental QALYs multiplied by the ceiling ratio (CR) of willingness to pay (WTP) per QALY minus the incremental costs.

> NMB = (QALYs * CR) – Costs

### Sensitivity Analysis

Both probabilistic and deterministic sensitivity analyses were conducted to explore the model’s uncertainty. The deterministic sensitivity analysis was conducted to investigate the impact of key assumptions and parameter values on the base-case analysis, including discount rate, time horizon, and treatment effect. The probabilistic sensitivity analyses were modeled through Monte Carlo simulation. Uncertainty in each of the underlying modeling parameters were characterized by assigning probability distribution to point estimates, and the model was run for 10,000 times for baseline estimate. Results were presented as plotted around point estimates of ICER on Incremental cost effectiveness plane.

## Results

The primary objective of the simulation model was to calculate the accumulated events of four diabetes-related complications over the lifetime of 2,931 patients with specific baseline characteristics and risk factors. As shown in Table 4.2, over the lifetime horizon of usual care patients, there were 206 heart failures, 242 strokes, 29 amputations, and 51 cases of blindness. In comparison, over the lifetime horizon of pharmacy-based intervention patients, 159, strokes, 19 heart failures, 24 amputations and 29 cases of blindness events could be averted according to our model. The model also predicted 155 fewer death associated with complications among intervention groups compared to usual care over the lifetime horizon. The cumulative cost was discounted at 3% per year. Over 50 years, the cumulative discounted cost for usual care patients was $30,159,963 CAD total or $10,289 CAD per patient. In comparison, the cumulative discounted cost for pharmacy-based intervention patients was only slightly higher, at $33,904,268 CAD total or $11,567 CAD per patient.

The pharmacy-based intervention was associated with 0.42 additional life-years and 0.32 additional QALYs per patient in comparison to usual care. Cumulatively, the pharmacy-based intervention was associated with 15,207 QALYs or 5.18 per patient, whereas usual care was associated with 14,254 QALYs or 4.8 per patient ***(Table 2)***.

**Table 2).** Result of Microsimulation Model.

|  | Usual care | Pharmacist Intervention | Difference |
| --- | --- | --- | --- |
| **Base case** |  |  |  |
| Accumulated number of heart failure | 206 | 187 | 19 |
| Accumulated number of strokes | 242 | 83 | 159 |
| Accumulated number of amputations | 29 | 5 | 24 |
| Accumulated number of blindness | 51 | 22 | 29 |
| Accumulated number of deaths associated with events | 401 | 246 | 155 |
| Quality Adjusted Life Years (QALYs) | 14,254 | 15,207 | 953 |
| QALYs per patients | 4.8 | 5.18 | 0.32 |
| Discounted Costs | \$30,159,973 | \$33,904,168 | \$3,744,195 |
| Discounted Costs per patients | \$10,289 | \$11,567 | \$1277 |
| Undiscounted Costs | \$40,846,595 | \$46,355,459 | \$5,508 |
| ICER (Discounted) | | | \$3928 |
| ICER (Undiscounted) | | | \$5779 |
| NMB | | | \$12,072 |
| **One-way sensitivity Analysis** |  |  |  |
| ICER (20 Years horizon) | | | \$2831 |
| ICER (Low discount rate) | | | \$4923 |
| ICER (Reduce treatment effect) | | | \$4580 |

Improvements in HbA1c, BMI, SBP and LDL expected to result from the pharmacy-based interventions reduced the number of diabetes-related complications over the lifetime horizon. This reduction led to the addition of 0.32 QALYs, and an ICER of $3,929 CAD per QALY compared to usual care.

Despite changes to the ICERs across the one-way sensitivity analyses, the pharmacy-based intervention remained cost-effective. When the lifetime horizon was shortened to 20 years, the ICER changed to $2,831.9 CAD. When the lifetime horizon was shortened to 20 years and a 1.5% discount was applied to costs and effects, ICER changed to $4,923. When the anticipated effect of the intervention on HbA1c, BMI, LDL, and SBP was lowered, ICER changed to $4,580 ***(Table 2)***.

Our probabilistic sensitivity analyses involved varying all of the parameters’ uncertainties at the same time using Monte Carlo simulation. Then, 10,000 samples of cost and QALYs were used for both the usual care and intervention groups ***(Figure 2)***. As shown in the cost-effectiveness acceptability curve plot, 92% of iterations remained within a cost-effectiveness threshold of $50,000 per QALY ***(Figure 3)***.

**Figure 2).**
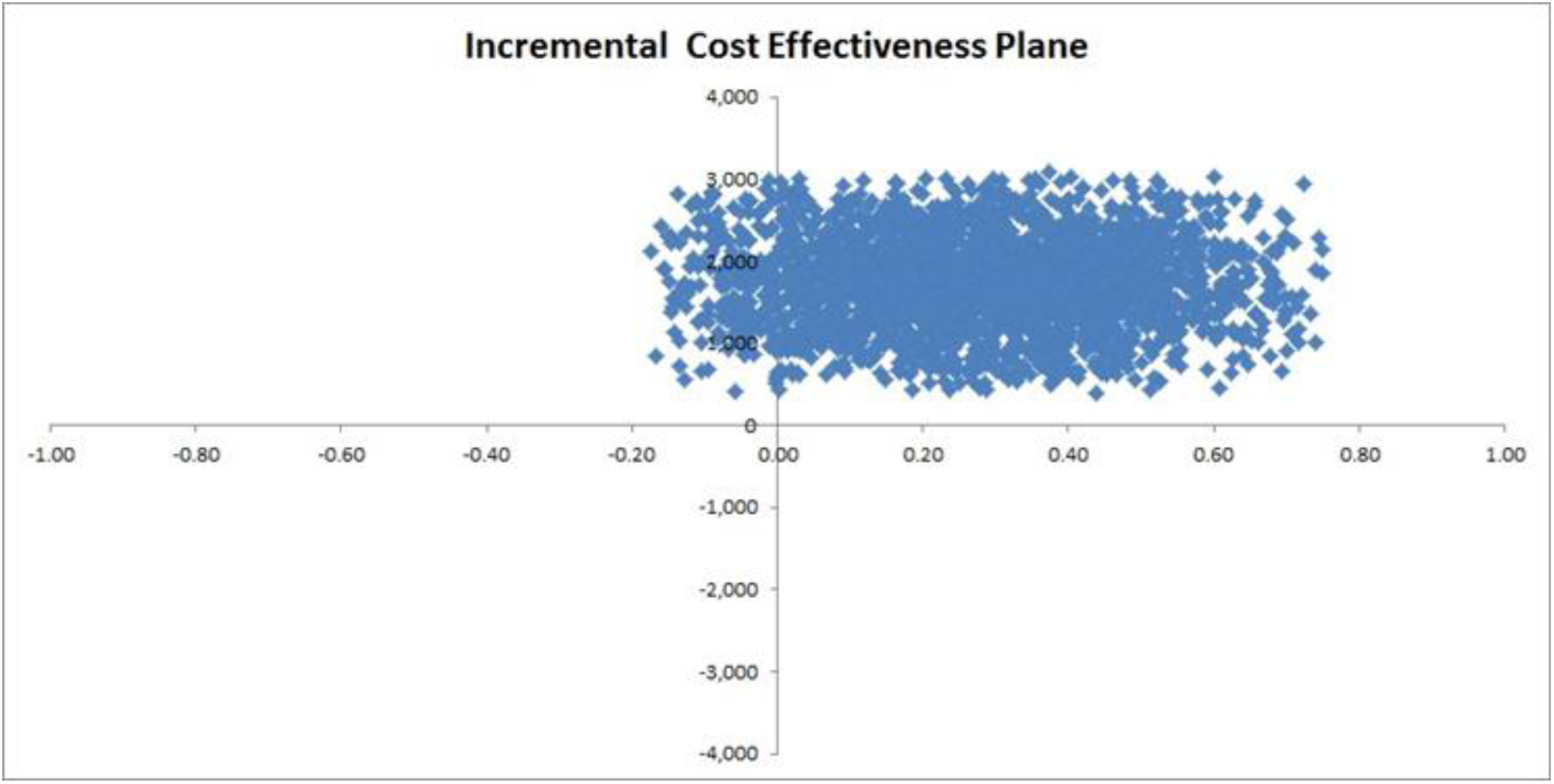
Incremental cost effectiveness plane.

**Figure 3).**
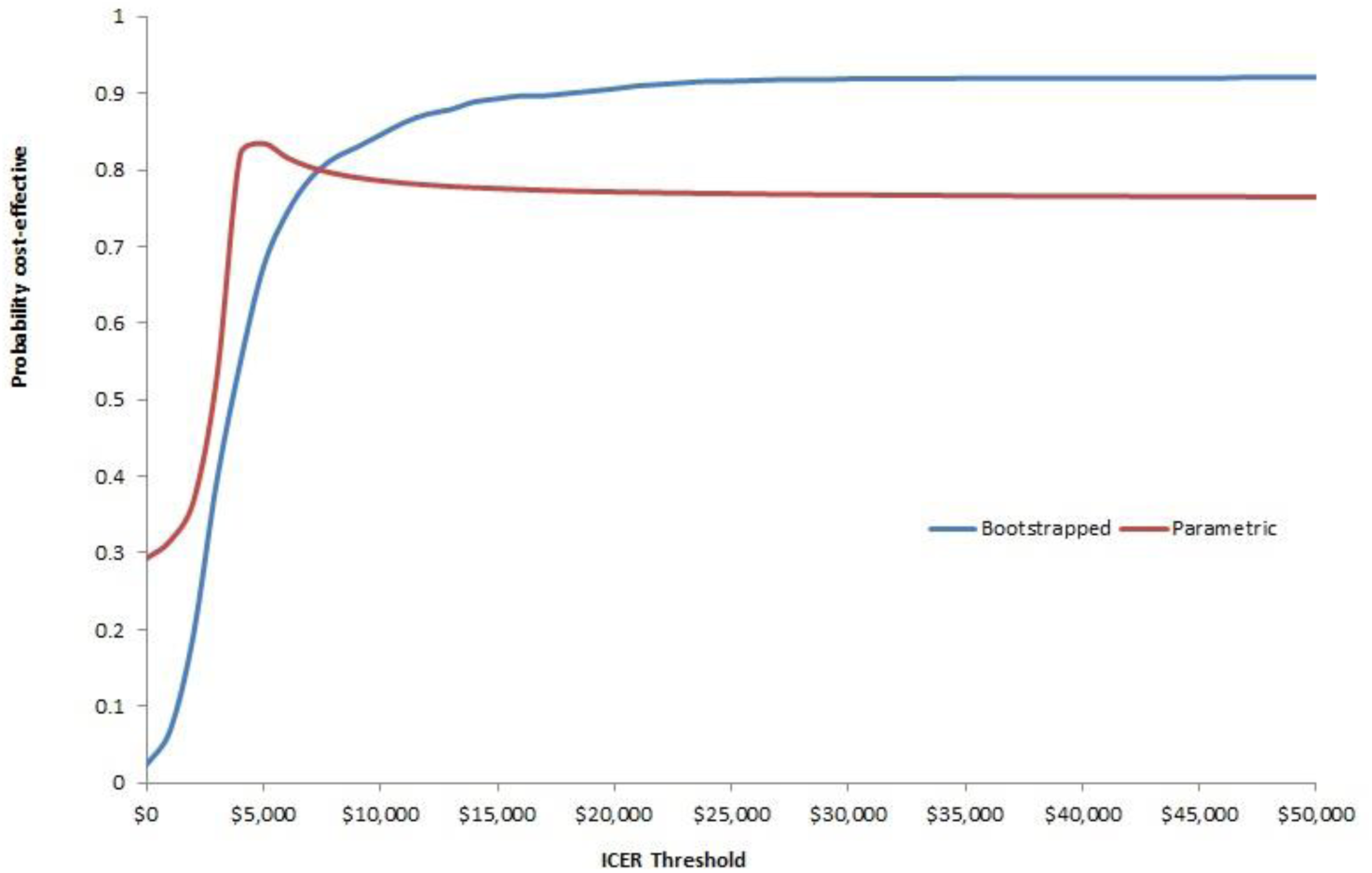
cost effectiveness acceptability curve.

## Discussion

Using a patient-level micro-simulation model, the future potential cost-effectiveness of pharmacy-based intervention in the control and management of four diabetes-related complications was analyzed. Across a 50-year lifetime horizon, the intervention proved to be a cost-effective strategy when compared to the usual care (status quo) diabetes patients across Canada receive. The ICER remained below the cost-effectiveness threshold across both deterministic and probabilistic sensitivity analyses. This suggests that the implementation of pharmacy-based interventions could yield consistent results. This consistency is encouraging, especially in the context of the Canadian health care system where pharmacists’ scope is impacted by the jurisdiction in which they practice (36-37).

The treatment effect of pharmacy-based interventions was assumed to be low in our analysis. The effect on HbA1c, BMI, SBP and LDL was set at 0.71, 0.20, 4.6, and 7.6 respectively. Based on these values, the intervention resulted in the addition of 670 more QALYs compared to status quo. Our analysis included a wide range of services that made up an integrated pharmacy-based intervention. However, the results of a recent meta-analysis suggest that diabetes education delivered by pharmacists coupled with pharmaceutical care maximizes the effectiveness of such services (28). HbA1c was lowered by 0.86 and SBP was lowered by 4.94 when this scheme was followed (28). When our model was adjusted in accordance with this specific type of pharmacy-based intervention, the result did not change. Further research must be conducted to corroborate these findings, but this result suggests that pharmacy-based intervention could be effective in even the most resource limited settings.

Our results did change, however, when the discount rate and time horizon were reduced. In these scenarios, the pharmacy-based intervention group gained 1,031 and 1,112 QALYs respectively. These results highlight that this interventions will operate across jurisdictions and across patient lifespans.

Our findings add to the literature and support previous economic evaluations of simulated community care programs for diabetes patients in Canada and around the world. Another Canadian patient-level microsimulation mode, the Ontario Diabetes Economic Model (ODEM), suggested that the incremental cost-effectiveness per QALY of a multidisciplinary management program was $5,203 over 10 years (22). Pharmacy-based interventions have also proven to be cost-effective in reducing risk factors, like hypertension, associated with diabetes-related complications in Canadian patients (29). This mirrors the reductions in risk factors reported in the present study. Further, a Markov cost-effectiveness model evaluated pharmacy-based interventions in Kaiser Permanente Northern California and results demonstrated saving of $6,364 over the lifetime of a single diabetes patient; prevent cardiovascular disease among diabetes patients; and, is less expensive and more effective than usual care over a 10-year time span (30). Again, these positive results compliment the findings from present study. By adding to this literature, provincial governments and health professionals are given a source from which to draw when creating evidence-based practices for the treatment of diabetes patients.

Additionally, our findings compliment previous economic evaluations conducted in real time using emerging patient data. Over 36-months, a randomized controlled clinical trial estimated the ICER per QALY of pharmaceutical care used to manage diabetes and hypertension among elderly patients (31). This analysis demonstrated that pharmaceutical care did not significantly increase the cost of direct health care, but did significantly improve health outcomes (31). More specifically, the ICER per QALY ($53.50) gained reflected favorably on its cost-effectiveness (31). In a North American context, the cost-effectiveness of pharmacy-led drug management education programs (DMEP) has been evaluated. Results from these evaluations suggest that DMEPs are cost-effective relative to usual care, and avert $39 USD per day spends on glycemic symptoms among diabetes patients (32). The results from our simulated model are consistent with findings from clinical trials, and operational education programs. This consistency reinforces the idea that pharmacy-based interventions could be an effective means to manage diabetes and its related complications.

Lastly, our findings do not vary even when compared to the results yielded by different modeling techniques. When a discrete-event simulation model was used, alternative treatment strategies, like pharmacy-based interventions, were associated with enhanced long-term health outcomes among diabetes patients (33). Similarly, when a Markov cohort analysis was conducted to assess the cost-effectiveness of Continuous Glucose Monitoring (CGM) technology compared to self-monitoring, CGM led to an expected improvement of 0.52 QALYs and was cost-effective in 70% of the Monte Carlo simulations (34). When a decision tree model was used to estimate the cost-effectiveness of pharmacy-based ophthalmology screenings compared to in-person examinations, the ICER was $314 CAD per additional case detected, and $73 CAD per additional case correctly diagnosed (35).

Also, in another study authors used a modified Sheffield T1D policy model to simulate T1D complication and estimate cost effectiveness of continuous Glucose monitoring in diabetes trial consist of 158 patients. Result of this analysis demonstrated that, continuous glucose monitoring led to an ICER of $98,000 and not only improve HbA1c controlling but also is cost –effective intervention in threshold of $100,000 in USA. Lastly an economic model was developed by Houle SK and et al (37) to estimate the effect of a pharmacist-based hypertension management program on economic burden of health care system. Results of this study determined that, this intervention could save $115 per patient for a program lasting one year (37).

## Conclusion

In conclusion, Canadian health policy makers should afford consideration to pharmacy-based interventions in the management of diabetes and its related complications. Through the expansion of education-based services by community pharmacies, there is the potential to reduce the incidence of diabetes-related complications, and death resulting from diabetes. This is an important avenue that should not be overlooked, especially because it offers a solution to the growing and complex problems caused by diabetes.

## Data Availability

all data are available upon request

## Acknowledgment

We thank Megan Steeves and Aydin Teyhouee for their invaluable cooperation.

## Appendix

**Figure A1).**
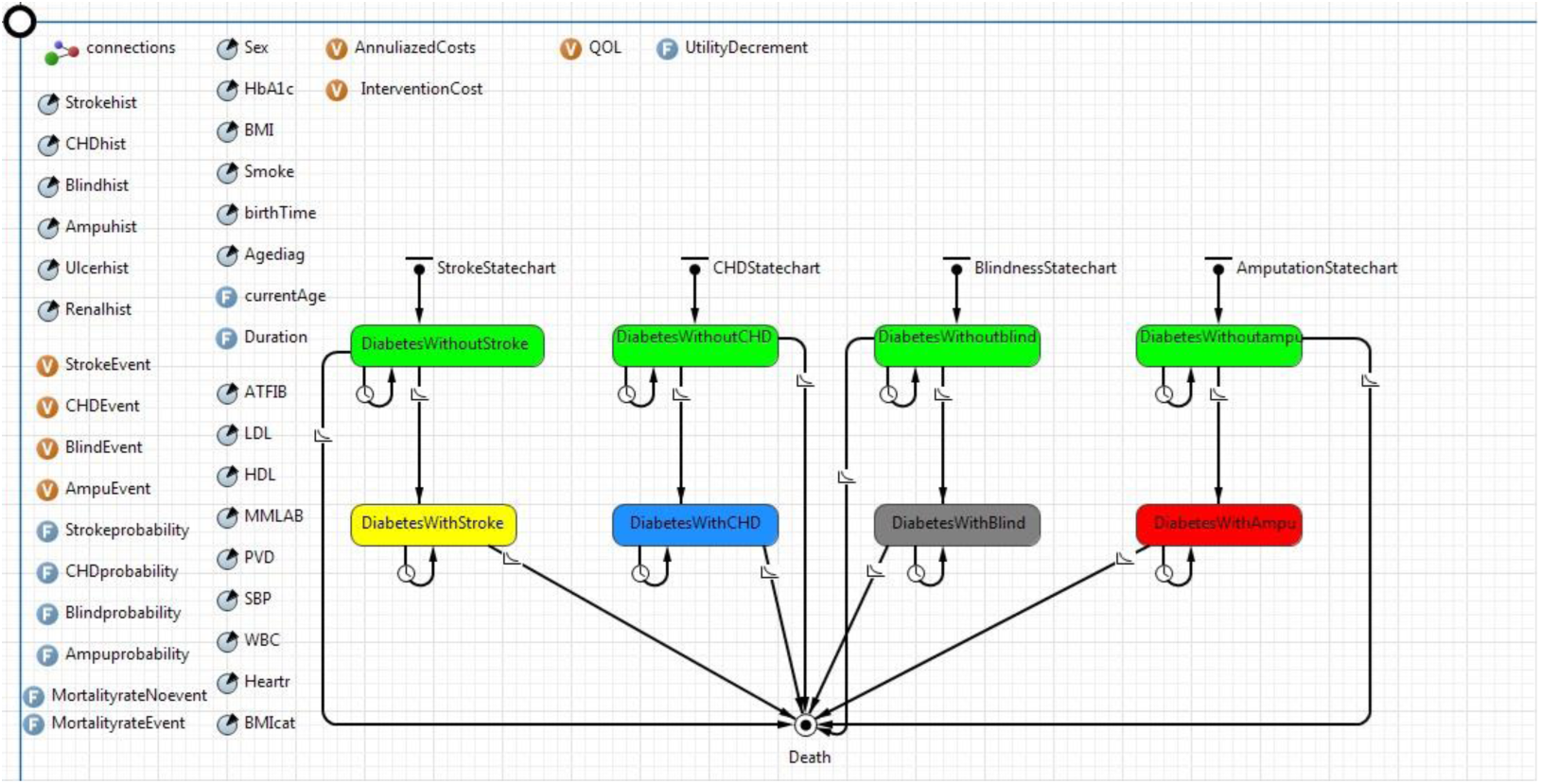
Agent-based model architectures in Anylogic.

**Figure A2).**
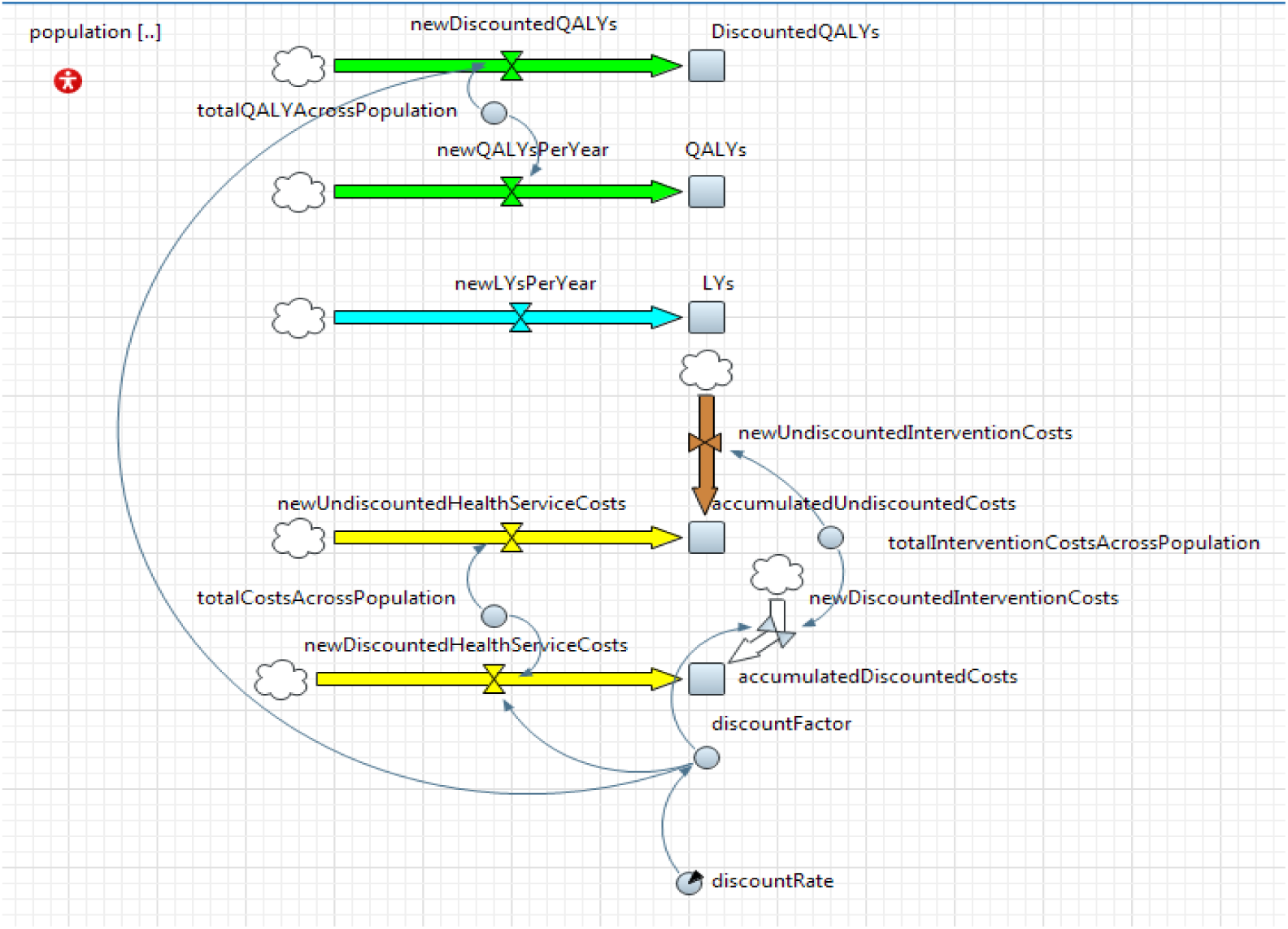
System dynamic model architectures in Anylogic.

### Appendix I) Estimating the progression from diabetes with no complication to diabetes-related complication state

**Table A1).** Risk equation for developing four diabetes-related complications based on UKPDS study.

| Risk equation for developing complications | Mean | Se | Functional form | Source |
| --- | --- | --- | --- | --- |
| **Stroke \*** |  |  |  |  |
| $\lambda$ | -13.053 | 1.41 | Weibull | Hayes AJ (2013) |
| P | 1.466 | 0.15 | Weibull | Hayes AJ (2013) |
| Age of Diagnoses | 0.666 | 0.01 | Weibull | Hayes AJ (2013) |
| Sex | -0.42 | 0.19 | Weibull | Hayes AJ (2013) |
| HbA1c | 0.092 | 0.05 | Weibull | Hayes AJ (2013) |
| Smoke | 0.331 | 0.21 | Weibull | Hayes AJ (2013) |
| Heart Failure history | 0.481 | 0.27 | Weibull | Hayes AJ (2013) |
| Amputation history | 1.09 | 0.47 | Weibull | Hayes AJ (2013) |
| LDL | 0.016 | 0.007 | Weibull | Hayes AJ (2013) |
| ATFIB | 1.467 | 0.39 | Weibull | Hayes AJ (2013) |
| MMALB | 0.42 | 0.19 | Weibull | Hayes AJ (2013) |
| SBP | 0.17 | 0.04 | Weibull | Hayes AJ (2013) |
| WBC | 0.04 | 0.01 | Weibull | Hayes AJ (2013) |
| **Heart Failure \*** |  |  |  |  |
| $\lambda$ | -12.332 | 1.6 | Weibull | Hayes AJ (2013) |
| P | 1.514 | 0.17 | Weibull | Hayes AJ (2013) |
| Age of Diagnoses | 0.068 | 0.01 | Weibull | Hayes AJ (2013) |
| BMI | 0.072 | 0.01 | Weibull | Hayes AJ (2013) |
| Amputation History | 0.0658 | 0.64 | Weibull | Hayes AJ (2013) |
| Ulcer foot History | 0.654 | 0.57 | Weibull | Hayes AJ (2013) |
| ATFIB | 1.562 | 0.48 | Weibull | Hayes AJ (2013) |
| LDL | 0.012 | 0.009 | Weibull | Hayes AJ (2013) |
| MMALB | 0.771 | 0.22 | Weibull | Hayes AJ (2013) |
| PVD | 0.479 | 0.26 | Weibull | Hayes AJ (2013) |
| **Amputation\*** |  |  |  |  |
| $\lambda$ | -14.844 | 2.36 | Weibull | Hayes AJ (2013) |
| P | 2.067 | 0.37 | Weibull | Hayes AJ (2013) |
| Age of Diagnoses | 0.023 | 0.02 | Weibull | Hayes AJ (2013) |
| Sex | -0.445 | 0.37 | Weibull | Hayes AJ (2013) |
| HbA1c | 0.248 | 0.08 | Weibull | Hayes AJ (2013) |
| Stroke History | 1.299 | 0.47 | Weibull | Hayes AJ (2013) |
| ATFIB | 1.088 | 0.78 | Weibull | Hayes AJ (2013) |
| HDL | -0.059 | 0.06 | Weibull | Hayes AJ (2013) |
| MMALB | 0.602 | 0.35 | Weibull | Hayes AJ (2013) |
| PVD | 1.01 | 0.37 | Weibull | Hayes AJ (2013) |
| SBP | 0.086 | 0.07 | Weibull | Hayes AJ (2013) |
| WBC | 0.04 | 0.01 | Weibull | Hayes AJ (2013) |
| Heart rate | 0.098 | 0.09 | Weibull | Hayes AJ (2013) |
| **Blindness** |  |  |  |  |
| $\lambda$ | -11.607 | 1.48 | Exponential | Hayes AJ (2013) |
| Age of Diagnoses | 0.047 | 0.01 | Exponential | Hayes AJ (2013) |
| HbA1c | 0.171 | 0.06 | Exponential | Hayes AJ (2013) |
| Heart Failure History | 0.841 | 0.54 | Exponential | Hayes AJ (2013) |
| Stroke History | 0.61 | 0.39 | Exponential | Hayes AJ (2013) |
| SBP | 0.068 | 0.06 | Exponential | Hayes AJ (2013) |
| WBC | 0.052 | 0.03 | Exponential | Hayes AJ (2013) |
| Heart rate | 0.08 | 0.05 | Exponential | Hayes AJ (2013) |

Following descriptions derived from the supplementary material of United Kingdom Prospective Diabetes model (UKPDS2) (1)

The Weibull model of UKPDS for heart failure in table D assumes a baseline hazard given by:

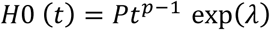

And in the proportional hazards model;

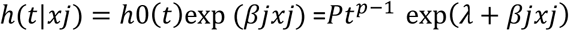

The parameters requiring calculation are λ, βj and p, which are given in Table D; time at risk (t) in the model is duration of diabetes. The unconditional probability of heart failure occurring between time t and t+1 can be estimated using the integrated hazard. The integrated hazard at time t is:

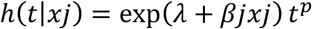

And the unconditional probability of heart failure in the interval t to t+1 is:

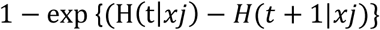

For example the calculation of the probability of heart failure in the current year for a one patient record in SLCDC with following characteristic (male, 70 years of age, with 8 years of diabetes, LDL 3.0 mmol/l, BMI of 32, eGFR 50, with microalbuminuria and a history of amputation) estimated as below:

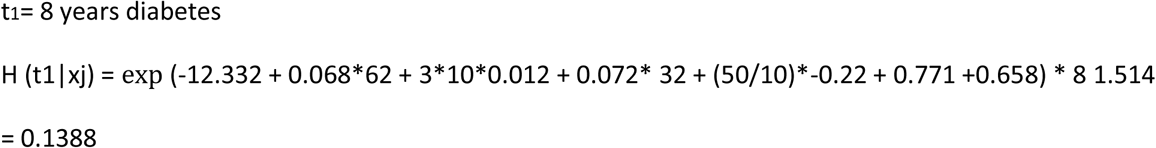

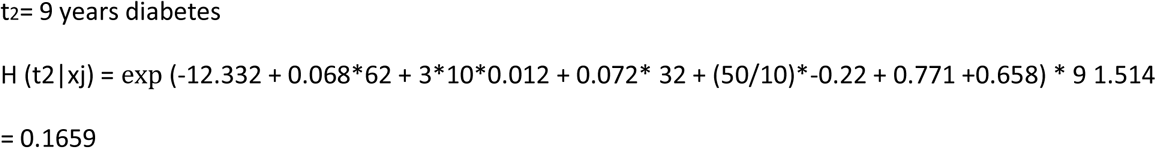

Probability of heart failure in current year= 1-exp (0.1388-0.1659) = 0.027

### Appendix II) Estimating the Probability of death in the 1st year of complication/s and no history of events

**Table A2).** Death equations based on UKPDS study.

| **Death equation (No History / No event)** | **Mean** | **Se** | **Functional form** | **Source** |
| --- | --- | --- | --- | --- |
| $\lambda$ | -10.908 | 0.58 | <u>Gompertz</u> | Hayes AJ (2013) |
| $\phi$ | 0.098 | 0.007 | <u>Gompertz</u> | Hayes AJ (2013) |
| Sex | -0.229 | 0.15 | <u>Gompertz</u> | Hayes AJ (2013) |
| Smoke | 0.379 | 0.17 | <u>Gompertz</u> | Hayes AJ (2013) |
| **Death equation (History/No event)** |  |  |  |  |
| $\lambda$ | -9.207 | 1.08 | <u>Gompertz</u> | Hayes AJ (2013) |
| $\phi$ | 0.073 | 0.01 | <u>Gompertz</u> | Hayes AJ (2013) |
| Smoke | 0.374 | 0.26 | <u>Gompertz</u> | Hayes AJ (2013) |
| Heart Failure History | 0.632 | 0.25 | <u>Gompertz</u> | Hayes AJ (2013) |
| Stroke History | 0.473 | 0.23 | <u>Gompertz</u> | Hayes AJ (2013) |
| Amputation history | 0.539 | 0.38 | <u>Gompertz</u> | Hayes AJ (2013) |
| BMI | -0.293 | 0.23 | <u>Gompertz</u> | Hayes AJ (2013) |
| MMALB | 0.348 | 0.20 | <u>Gompertz</u> | Hayes AJ (2013) |
| WBC | 0.048 | 0.02 | <u>Gompertz</u> | Hayes AJ (2013) |
| Renal History | 1.15 | 0.38 | <u>Gompertz</u> | Hayes AJ (2013) |
| **Death equation (First Year of Event)** |  |  |  |  |
| $\lambda$ | -6.916 | 1.15 | Logistic | Hayes AJ (2013) |
| Age | 0.058 | 0.01 | Logistic | Hayes AJ (2013) |
| Duration | 0.042 | 0.01 | Logistic | Hayes AJ (2013) |
| Smoke | 0.444 | 0.22 | Logistic | Hayes AJ (2013) |
| Heart Failure Event | 1.309 | 0.30 | Logistic | Hayes AJ (2013) |
| Stroke Event | 0.547 | 0.34 | Logistic | Hayes AJ (2013) |
| Amputation Event | -0.734 | 0.62 | Logistic | Hayes AJ (2013) |
| Heart rate | 0.124 | 0.06 | Logistic | Hayes AJ (2013) |
| PVD | 0.367 | 0.25 | Logistic | Hayes AJ (2013) |
| **Death equation (Subsequent Year of Event)** |  |  |  |  |
| $\lambda$ | -4.868 | 1.62 | Logistic | Hayes AJ (2013) |
| Age | 0.05 | 0.01 | Logistic | Hayes AJ (2013) |
| Heart Failure History | -0.507 | 0.37 | Logistic | Hayes AJ (2013) |
| Heart Failure Event | 0.583 | 0.47 | Logistic | Hayes AJ (2013) |
| Stroke History | 0.44 | 0.35 | Logistic | Hayes AJ (2013) |
| Stroke Event | -0.619 | 0.47 | Logistic | Hayes AJ (2013) |
| Amputation History | 0.753 | 0.58 | Logistic | Hayes AJ (2013) |
| Amputation Event | -1.267 | 0.66 | Logistic | Hayes AJ (2013) |
| Renal disease History | 0.089 | 0.07 | Logistic | Hayes AJ (2013) |
| WBC | 0.584 | 0.58 | Logistic | Hayes AJ (2013) |
| PVD | 0.352 | 0.34 | Logistic | Hayes AJ (2013) |
| ATFIB | 1.0817 | 0.76 | Logistic | Hayes AJ (2013) |
| HDL | 0.068 | 0.05 | Logistic | Hayes AJ (2013) |

Following descriptions derived from the supplementary material of United Kingdom Prospective Diabetes model (UKPDS2) (1)

Probability of death in the 1st year of complication/s and no history of events:

Logistic regression was used to model mortality in the year of an event. The probability of survival is given by;

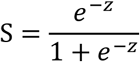

Where

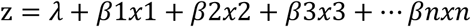

#### Current year MI

As an example, using the coefficients in Table E, for a one patient record in SLCDC with following characteristic (70 year-old male smoker with 12 years of diabetes, heart rate 80 bpm, has an MI but has no history of other events)

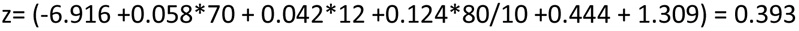

Probability of survival = exp (−0.393)/ (1+ exp (−0.393)) =0.403

Probability of death in current year = 0.597

#### Current year heart failure

Similarly, the same person with heart failure

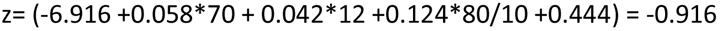

Probability of survival = exp (0.916)/(1+ exp (0.916)) =0.714

Probability of death in current year = 0.286

Also Gompertz regression model was used, in which the hazard of death increases exponentially with age. The Gompertz model assumes a baseline hazard given by:

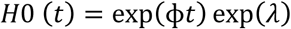

and in the proportional hazards model

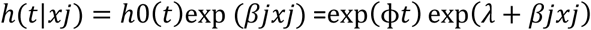

The parameters required are λ, βj and ϕ, which are given in table E; time at risk (t) in the model is current age. The probability of death occurring between time t and t+1 can be estimated using the integrated hazard. The integrated hazard at time t is:

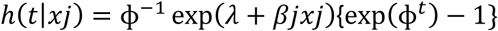

And the unconditional probability of death in the interval t to t+1 is:

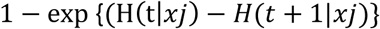

As an example, using the coefficients in Table E, for a patient record in SLCDC with following characteristics (a 70 year-old male smoker with 12 years of diabetes, who is overweight, has a white blood cell count of 6×106/ml and a history of heart failure but no events in the current year)

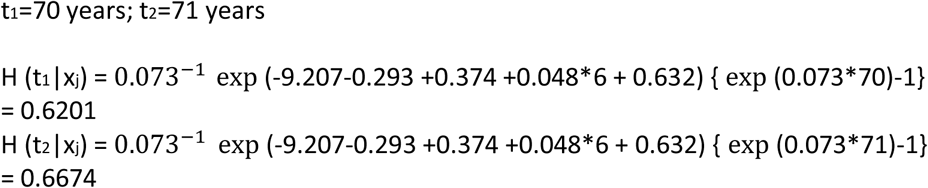

Probability of death between age 70 and 71 years = 1-exp (0.6201-0.6674) =0.0462

